# Genomic foundation model-derived disruption profiling links somatic mutations to cancer biology and clinical outcomes

**DOI:** 10.64898/2026.09.15.26363174

**Authors:** Akshatha Nayak, Tae-Rim Lee, Vikram Agarwal, Ilias Georgakopoulos-Soares

## Abstract

Cancer genomics has concentrated on individual mutations, overlooking whether somatic mutations can accumulate to produce partial, gene-level disruption with biological and clinical consequences. Sequence-to-function models can quantify these effects directly from DNA sequence. Here, we use AlphaGenome and AlphaMissense to quantify the disruption imposed by somatic mutations across 8,800 patients and 33 cancer types from The Cancer Genome Atlas. At the individual-variant level, recurrent hotspot mutations showed substantially larger predicted protein-level effects, whereas non-hotspot mutations exhibited larger regulatory effects across most cancer types. We then aggregated the variant-level predictions to construct patient-gene disruption profiles capturing transcriptional activity, chromatin accessibility, transcription factor binding, and splicing. These profiles were gene- and modality-specific, and reflected tissue of origin, cancer type, and microsatellite-instability status while retaining information beyond tumor mutational burden. Among patients lacking recurrent hotspot mutations in a given cancer gene, higher predicted disruption was associated with overall survival, with the strongest and most consistent signal observed for chromatin accessibility. In an independent treatment-annotated cohort, gene-level disruption was also associated with survival within treatment-defined subgroups. Together, these findings show that recurrent hotspots are enriched for strong predicted protein-level effects, whereas regulatory consequences are distributed more broadly across other variants, supporting a continuous, multidimensional view of cancer-gene perturbation beyond discrete drivers.

## Introduction

Cancer-associated somatic mutational analyses have primarily focused either on the identification of driver genes and driver mutations (Vogelstein et al. 2013; Bailey et al. 2018) or on aggregate mutational burden and mutational signature analyses (Rizvi et al. 2015; Alexandrov et al. 2013; Sha et al. 2020). However, whether somatic mutations can accumulate within a gene to progressively inactivate it, rather than acting as discrete, independent events, remains largely unexplored. We recently showed that joint analysis of somatic mutations within a cancer gene can reveal cumulative functional effects not captured by single mutation analyses, supporting a continuous model of gene disruption rather than the conventional binary view (Nayak et al. 2026).

Extending this framework requires a way to quantify how strongly a tumor’s somatic mutations perturb the function of a cancer gene. Recurrent protein-coding mutations are commonly prioritized because they have substantial functional and clinical impact, including effects on drug sensitivity, resistance, and therapeutic response (Chang et al. 2016). However, coding hotspot-based approaches are less sensitive to rare, dispersed, and cumulative regulatory effects, which may be mediated through different mechanisms such as chromatin state, transcriptional regulation, or RNA processing (Cao et al. 2020; Rheinbay et al. 2020). Somatic mutations classified as passengers may therefore have limited predicted protein-level pathogenicity while still in certain cases perturbing gene function. Directly comparing protein-oriented and regulatory sequence-model predictions can help determine whether these two classes of models capture distinct components of cancer development.

Sequence-to-function models trained on large-scale genomic data, such as Enformer (Avsec et al. 2021), Borzoi (Linder et al. 2025), and AlphaGenome (Avsec et al. 2026), predict the effects of sequence variation on chromatin accessibility, gene expression, and splicing across tissues and cellular contexts. Because these models operate directly on DNA and output multiple regulatory readouts, they can score the functional impact of a variant without relying on pre-defined, coding-centric features, capturing regulatory and non-coding effects that conventional driver analyses miss. These models learn rich, position-specific representations of sequence variation, yet whether they capture biologically and clinically meaningful properties of tumorigenesis remains largely untested.

Here we use AlphaGenome (Avsec et al. 2026) to quantify the regulatory disruption that somatic variants impose on cancer genes across 8,800 patients and 33 cancer types from The Cancer Genome Atlas (TCGA), scoring each gene across seven regulatory modalities spanning transcription, chromatin accessibility, and RNA processing. We first compare individual-variant effects predicted by AlphaGenome with protein-level pathogenicity scores from AlphaMissense (Cheng et al. 2023), testing whether recurrent cancer hotspots exhibit distinct protein and regulatory-effect profiles from passenger mutations. We then aggregated AlphaGenome predictions across all somatic mutations assigned to each patient-gene pair to generate gene-level disruption scores for each regulatory output, including chromatin accessibility, transcription factor binding, transcriptional output, and RNA splicing. We show that these disruption scores form gene- and modality-specific profiles that distinguish cancer genes, and that patient-level disruption features recapitulate known biological structure, including tissue of origin, cancer type, and microsatellite-instability status, while providing information beyond tumour mutational burden. We further find that predicted disruption is clinically informative. In patients without recurrent hotspot mutations in a given cancer gene, higher passenger-derived regulatory disruption was associated with survival, with the strongest association observed for chromatin accessibility. Gene-level disruption is likewise associated with treatment response in an independent cohort of 570 whole-genome sequenced tumor samples (Pleasance et al. 2020). Together, these analyses distinguish the strong protein-level effects enriched among recurrent hotspots from the more broadly distributed regulatory effects of passenger variation and support a continuous, multidimensional view of cancer-gene perturbation beyond conventional driver and burden measures.

## Results

Somatic mutations are often prioritized based on their predicted effects on protein sequence, an approach that overlooks variants that act by disrupting non-coding mechanisms of chromatin accessibility, transcriptional regulation, and splicing. To capture these effects, we used AlphaGenome (Avsec et al. 2026), a deep learning model that predicts regulatory activity directly from DNA sequence.

We first selected a representative patient-gene pair with high AlphaGenome-predicted RNA-seq disruption and multiple somatic mutations in the gene. In this *NRAS* gene from a Colorectal Cancer (TCGA-COAD) patient with ten somatic mutations, the patient-mutated sequence produced a distinct predicted RNA-seq profile relative to the reference sequence (**Figure 1A**). Several regions showed divergence between the reference and patient-mutated predictions, with most exonic regions showing increased predicted RNA-seq signal in the altered sequence. Comparison with observed GTEx and patient RNA-seq signal showed broadly concordant directionality across many regions, supporting the relevance of the predicted disruption.

**Figure 1.**
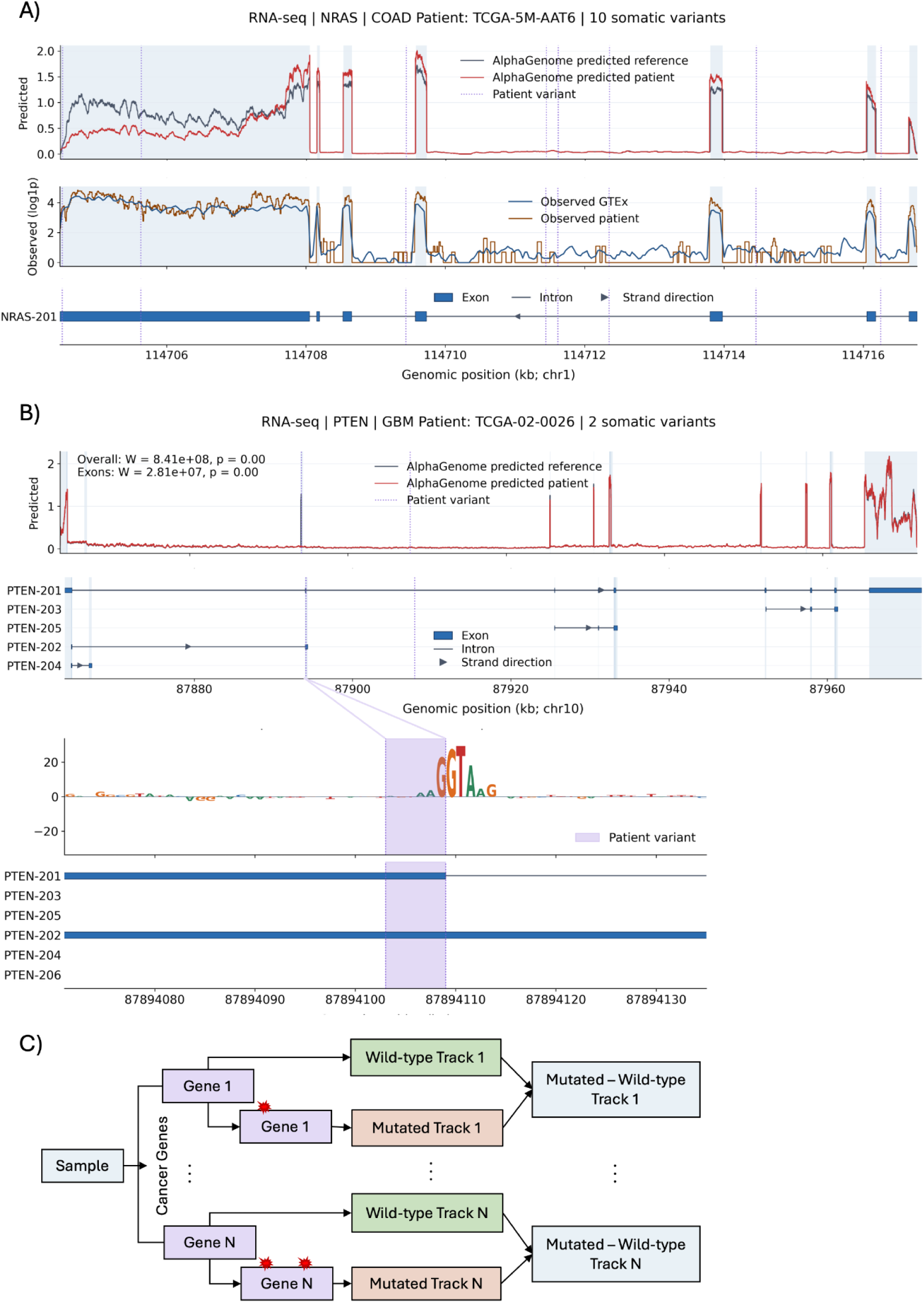
Predicting patient-specific regulatory disruption in cancer genes with AlphaGenome. **(A)** AlphaGenome-predicted RNA-seq signal for the NRAS locus in a COAD patient with 10 somatic variants. Predicted reference and patient-mutated tracks are shown alongside observed GTEx and patient RNA-seq signal, with dashed vertical lines marking patient variants and NRAS transcript structure shown below. **(B)** AlphaGenome-predicted RNA-seq signal for the PTEN locus in a GBM patient with two somatic variants. Predicted reference and patient-mutated tracks are shown with PTEN transcript isoforms below. The zoomed panel shows in-silico mutagenesis attribution for the splice-region deletion 10:87894104–87894109 (AGTAAG>A), with model importance concentrated at the disrupted canonical splice-donor motif. **(C)** Schematic showing that AlphaGenome is run on wild-type and mutated sequences, and the per-track difference (Mutated − Wild-type) quantifies predicted regulatory disruption.

We next examined a *PTEN* gene from a Glioblastoma (TCGA-GBM) patient with two somatic variants to illustrate how AlphaGenome predictions can localize somatic mutation effects to specific sequence features (**Figure 1B**). The patient-mutated sequence produced a distinct predicted RNA-seq profile relative to the reference sequence across *PTEN* transcript isoforms. *In-silico* mutagenesis (ISM) analysis further localized the predicted effect of a splice-region deletion, 10:87894104-87894109 (AGTAAG>A), to the disrupted canonical splice-donor motif. This example shows that AlphaGenome-predicted regulatory disruption can be traced back to interpretable sequence determinants, linking an individual somatic variant to altered transcript-level predictions.

Building on these examples, we used the reference-versus-alternate comparison to define patient-specific gene disruption (**Figure 1C**). For each patient, we scored every cancer gene by the per-track difference between mutated and reference predictions (Mutated - Wild-type) across seven regulatory modalities spanning transcription, chromatin accessibility, and RNA processing, yielding a patient-by-gene disruption profile. Applying this across 8,800 patients and 33 cancer types from the TCGA (The Cancer Genome Atlas) dataset produced the data analyzed in the succeeding sections.

### Hotspots show contrasting protein-level and regulatory effects to passenger mutations

We first compared the predicted effects of recurrent hotspot and passenger mutations at the individual somatic mutation level using AlphaMissense (Cheng et al. 2023) and AlphaGenome (Avsec et al. 2026). Across most cancer types, hotspot variants showed substantially greater AlphaMissense effects than passenger variants (adjusted q-value < 0.001 for all groups; **Figure 2A**). This pattern was observed consistently across both tumor-suppressor genes and oncogenes, with large differences in gene-normalized AlphaMissense scores between hotspot and passenger variants (TSGs: d = 1.75, q < 0.001; OGs: d = 1.17, q < 0.001; **Figure 2B**). These results are consistent with recurrent hotspots being enriched for mutations with strong predicted effects on protein sequence and function. In contrast, AlphaGenome-derived functional effects of passenger mutations showed the opposite overall pattern. Passenger mutations had greater predicted regulatory-effect magnitudes than hotspot variants across most cancer types, although the magnitude and significance of this difference varied among cohorts (**Figure 2A**). Passenger mutations retained higher AlphaGenome effects in both tumor-suppressor genes and oncogenes, with a larger difference among oncogenes (TSGs: d = −0.11, q < 0.001; OGs: d = −0.66, q < 0.001; **Figure 2B**). Thus, while hotspot mutations were strongly enriched for predicted protein-level pathogenicity, predicted regulatory effects were more broadly distributed among passenger mutations.

**Figure 2.**
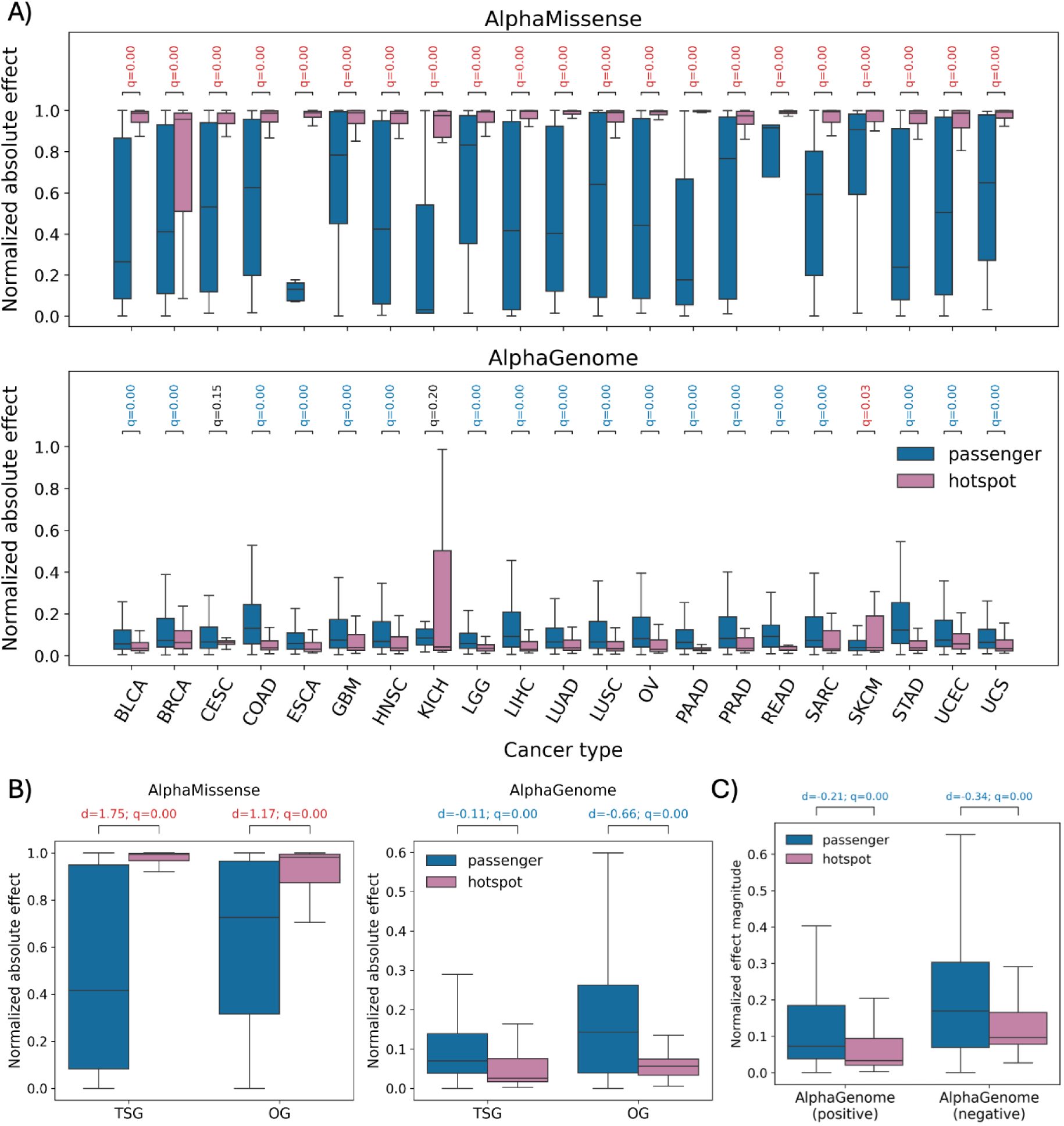
Hotspot and passenger variants show contrasting predicted protein-level and regulatory effects. **(A)** Distributions of per-variant normalized absolute effect scores for passenger and recurrent hotspot variants in top 10 pancancer driver genes, stratified by cancer types. The upper panel shows AlphaMissense scores, and the lower panel shows AlphaGenome-derived regulatory effect scores. **(B)** Normalized AlphaMissense and AlphaGenome effect scores stratified by tumor-suppressor genes (TSGs) and oncogenes (OGs). **(C)** AlphaGenome effects separated by predicted direction of change. Boxplots represent the median and interquartile range, with whiskers extending to 1.5 times the interquartile range. Brackets indicate comparisons between passenger and hotspot variants within each group. In all panels, statistical significance was assessed using a two-sided Mann–Whitney U test, and labels show Benjamini–Hochberg-adjusted q-values, with text color indicating the group with the higher median effect. Reported values above brackets indicate effect size d (in B, C) and BH-adjusted q-value. Scores are normalized within genes and cancer types for all panels.

We next separated AlphaGenome predictions according to the direction of the predicted regulatory change. Passenger variants showed greater effect magnitudes than hotspot variants among both mutations with positive predicted effects (d = −0.21, q < 0.001) and those with negative predicted effects (d = −0.34, q < 0.001; **Figure 2C**). Together, these findings indicate that recurrent hotspots preferentially capture mutations with strong protein-level consequences, whereas mutations classified as passengers can nevertheless produce substantial predicted regulatory perturbations in either direction. This contrast motivates evaluating somatic variation using regulatory sequence models in addition to conventional coding-centric pathogenicity scores.

### Regulatory and coding disruption reveal distinct gene- and tissue-specific changes

To determine whether these variant-level effects converge on broader patterns of gene disruption, we summarized AlphaMissense and AlphaGenome scores at the gene level across multiple cancer contexts. Specifically, we examined whether AlphaGenome-derived regulatory disruption and AlphaMissense-derived coding disruption produce similar or distinct gene-level profiles across tissues. Within each tissue or cancer context, disruption scores were normalized across modalities, adjusted for tumor mutational burden and gene length, and converted to percentiles, allowing genes to be compared relative to other cancer genes in the same context.

AlphaGenome disruption profiles revealed substantial tissue- and gene-specific heterogeneity (**Figure 3A**). Several genes, including *ARID1A*, *IDH1*, *KMT2C*, and *KMT2D*, showed high predicted regulatory disruption across multiple tissue contexts, whereas others displayed more restricted patterns. These differences suggest that the regulatory effects of somatic mutations are not uniform across cancer genes, but instead depend on both the affected gene and the tissue-specific regulatory landscape in which the variants occur.

**Figure 3.**
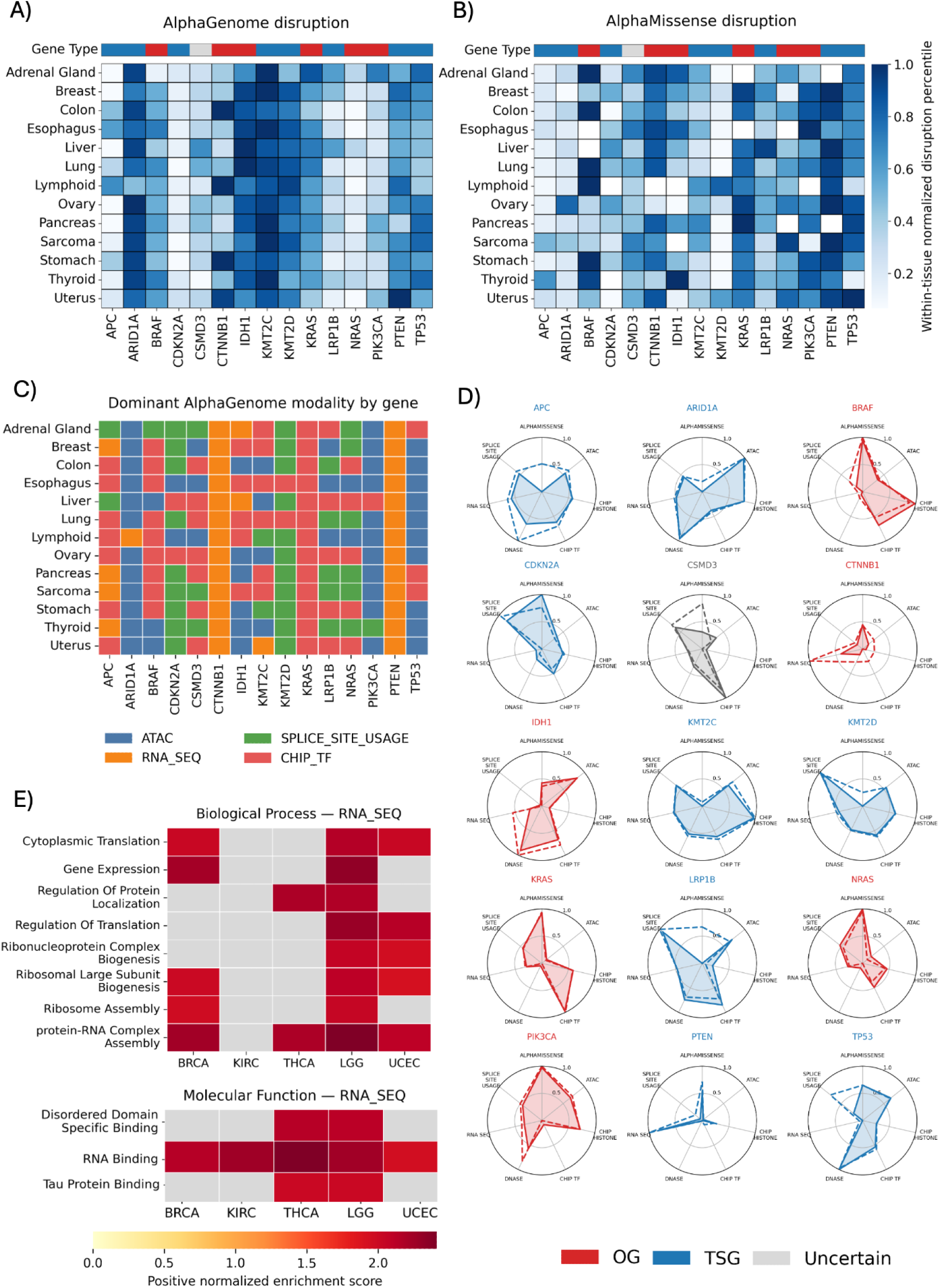
AlphaMissense and AlphaGenome reveal distinct tissue- and gene-specific patterns of somatic disruption. **(A)** Heatmap of within-tissue normalized AlphaGenome disruption percentiles across cancer genes and tissue contexts. Rows represent tissue or cancer contexts, columns represent genes, and the top annotation indicates gene class as oncogene, tumor suppressor, or uncertain/context-dependent. **(B)** Heatmap of within-tissue normalized AlphaMissense disruption percentiles for the same genes and tissue contexts, enabling comparison between regulatory disruption and coding pathogenicity patterns. **(C)** Dominant AlphaGenome output modality for each gene and tissue context, defined as the modality with the highest normalized disruption score. Colors indicate whether disruption is primarily captured by ATAC, RNA-seq, splice-site usage, or ChIP-TF outputs. **(D)** Radial plots showing modality-specific disruption profiles for top 15 pancancer genes. Solid and dashed lines show the median and mean, respectively, of patient-level disruption across AlphaMissense and AlphaGenome modalities, and background color denotes gene type. **(E)** GSEA of genes prioritized by RNA-seq disruption across cancer types. Genes were ranked by median absolute normalized RNA-seq disruption across patient observations, and significantly positively enriched Gene Ontology Biological Process and Molecular Function terms are shown. Positive enrichment indicates gene sets preferentially represented among genes with the strongest predicted RNA-seq disruption.

AlphaMissense disruption showed a partially distinct pattern from AlphaGenome disruption (**Figure 3B**). Some genes showed high disruption in both models, whereas others differed between the two approaches, indicating that coding pathogenicity and regulatory disruption capture complementary dimensions of somatic variant impact. This contrast extends the variant-level findings from Figure 2: recurrent hotspots are more strongly associated with protein-level pathogenicity, but aggregate gene-level disruption includes regulatory effects that are not captured by coding-centric scores alone.

We next examined which AlphaGenome output modality contributed most strongly to each gene-tissue disruption profile. The dominant-modality map showed that the leading regulatory signal varied substantially across genes and tissue contexts (**Figure 3C**). Some gene-tissue pairs were dominated by ATAC, consistent with altered chromatin accessibility, whereas others were dominated by RNA-seq, splice-site usage, or ChIP-TF outputs. Thus, AlphaGenome-derived disruption does not reflect a single dominant measure of variant impact, but instead resolves somatic regulatory perturbation into distinct functional layers, including accessibility, transcriptional output, splicing, and transcription factor binding. Radial plots of the top 15 pan-cancer genes further illustrated gene-specific disruption signatures across AlphaMissense and AlphaGenome modalities (**Figure 3D**). Some genes showed broad multimodal disruption, while others were dominated by a smaller number of modalities. Additionally, the close agreement between the mean and median profiles for most genes indicated that these patterns were generally not driven by a small number of extreme observations. These profiles provide an interpretable view of how somatic variation can perturb cancer genes through both protein-coding and regulatory mechanisms.

Finally, to test whether genes prioritized by AlphaGenome RNA-seq disruption represented coherent biological programs, we performed ranked GSEA using genes ordered by median absolute normalized RNA-seq disruption across the top five cancer types by sample size (**Figure 3E**). Genes with high RNA-seq disruption were positively enriched for RNA-associated and translation-related processes, including gene expression, cytoplasmic translation, ribosome assembly, ribonucleoprotein complex biogenesis, and protein-RNA complex assembly. Molecular function enrichment similarly highlighted RNA binding across all selected cancer types. These results support the interpretation that AlphaGenome RNA-seq disruption captures transcript-level perturbation in genes involved in RNA regulation, translation, and protein-RNA complex biology across multiple cancer types.

Together, these analyses show that the regulatory effects observed among passenger mutations at the variant level aggregate into structured, tissue-specific gene disruption profiles. AlphaMissense captures coding pathogenicity, whereas AlphaGenome resolves regulatory disruption into distinct modality-specific effects. This provides a mechanistic basis for evaluating somatic mutations using regulatory foundation models alongside conventional protein-centric pathogenicity scores.

### Disruption features recapitulate cancer identity and molecular subtype

We next asked whether patient-level disruption profiles capture known biological structure. Representing patients by their AlphaGenome disruption features and projecting with t-SNE, samples clustered by tissue of origin (pseudo-F=1132.070, p=0.001; **Figure 4A**) and by cancer type (pseudo-F=1676.180, p=0.001; **Figure 4B**), with thyroid (THCA), low-grade glioma (LGG), and endometrial (UCEC) tumors forming well-separated clusters. Thus, the regulatory disruption imposed by a tumor’s somatic mutations carries information about its tissue and disease context. Disruption features also separated tumors by microsatellite-instability (MSI) status within individual cancer types (**Figure 4C**). MSI-high tumors formed distinct clusters in stomach (STAD; pseudo-F=406.621, p=0.001) and endometrial (UCEC; pseudo-F=388.593, p=0.001) cancers, and were partially separated in colorectal cancer (COAD/READ; pseudo-F=161.449, p=0.001), indicating that disruption profiles reflect this clinically important molecular subtype.

**Figure 4:**
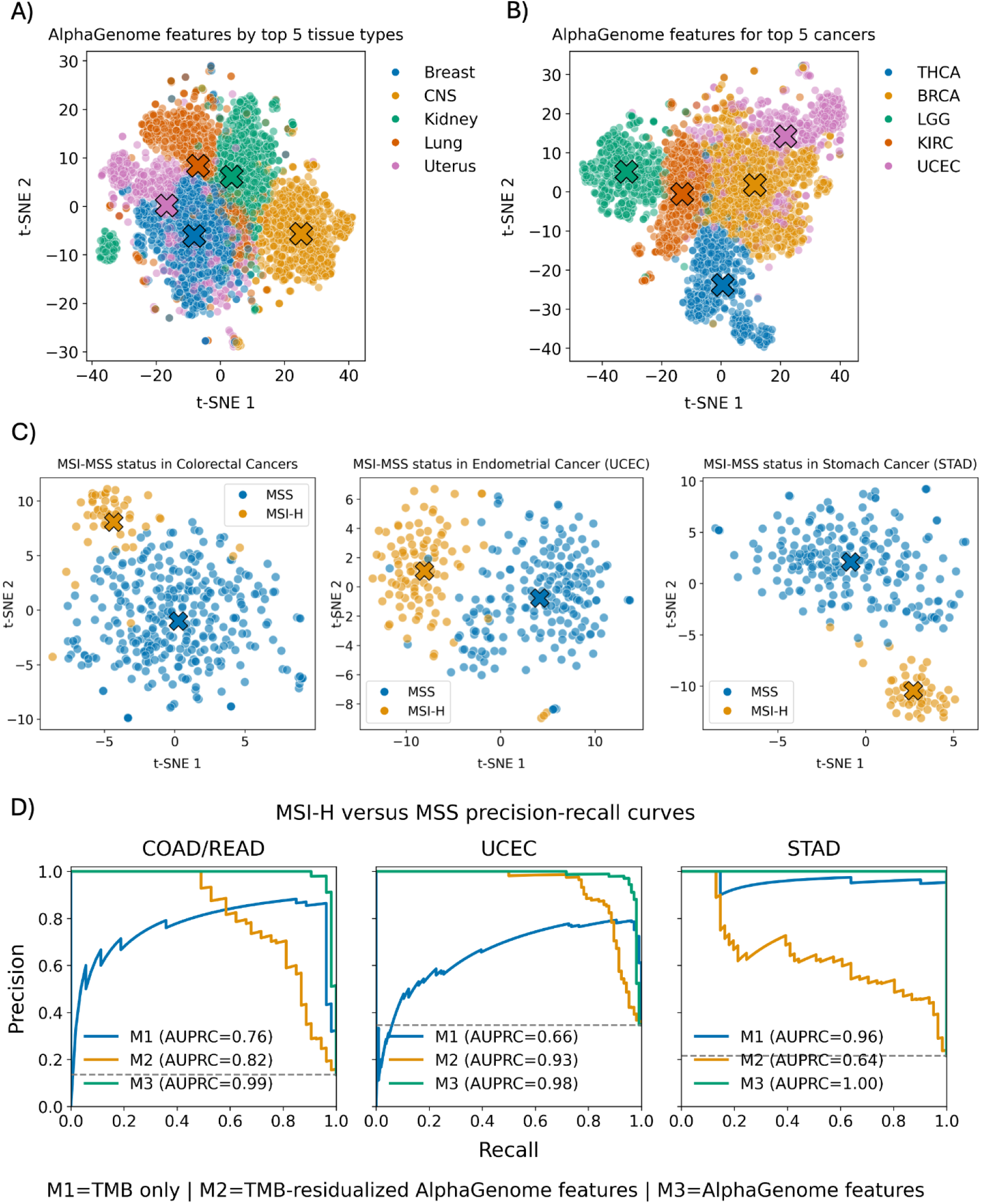
AlphaGenome-derived gene disruption scores capture cancer-specific and molecular subtype signatures. **(A)** tSNE plot clustering by tissue using AlphaGenome-derived disruption scores. **(B)** tSNE plot clustering by cancer subtypes (TCGA cohorts) using AlphaGenome-derived disruption scores. **(C)** tSNE plot clustering by MSI/MSS Status using AlphaGenome-derived disruption scores. The ’X’ markers indicate the centroids of the clusters. **(D)** Precision-Recall curves for MSI/MSS classifier models trained on TMB-only (Model M1), TMB-residualized AlphaGenome features (Model M2), and AlphaGenome features (Model M3).

Because MSI-high (MSI-H) tumors often carry elevated mutational burden, we tested whether the observed MSI-H/MSS separation was driven primarily by tumor mutational burden (TMB). We compared three MSI classifiers using precision–recall analysis: TMB alone (M1), TMB-residualized AlphaGenome features (M2), and full AlphaGenome features (M3) (**Figure 4D**). TMB alone showed moderate to strong but variable performance across cancer types, with AUPRC values of 0.76 in COAD/READ, 0.66 in UCEC, and 0.96 in STAD, consistent with the known relationship between MSI-H status and hypermutation. In contrast, the full AlphaGenome model achieved near-perfect classification across all three cancer types, with AUPRC values of 0.99 in COAD/READ, 0.98 in UCEC, and 1.00 in STAD. Importantly, TMB-residualized AlphaGenome features remained highly discriminative in COAD/READ and UCEC, with AUPRC values of 0.82 and 0.93, respectively, indicating that AlphaGenome disruption profiles retain MSI-relevant signal beyond overall mutational burden. In STAD, residualized AlphaGenome features performed less well than TMB alone, suggesting that MSI-associated AlphaGenome signal in this cancer type may be more closely coupled to mutation burden.

Together, these results show that AlphaGenome-derived disruption profiles recapitulate both cancer identity and MSI-associated molecular subtype structure. The improvement over TMB alone, particularly in COAD/READ and UCEC, supports the conclusion that regulatory disruption profiles capture subtype-relevant information that is not fully reducible to mutational burden.

### Predicted regulatory disruption from passenger mutations is associated with survival

Having shown that disruption profiles capture biological identity, we asked whether they carry prognostic information. We focused on patients lacking recurrent hotspot mutations in a given cancer gene, so that any signal would derive from passenger mutations rather than known drivers. For each gene, we residualized per-gene normalized disruption scores on tumor mutational burden (TMB), retained patients with non-zero disruption, and split them at the median into low- and high-disruption groups. Cox proportional-hazards models were then used to estimate hazard ratios for overall survival relative to patients with no predicted disruption and no hotspot mutation in that gene, including age and cancer type as covariates (**Figure 5**).

**Figure 5.**
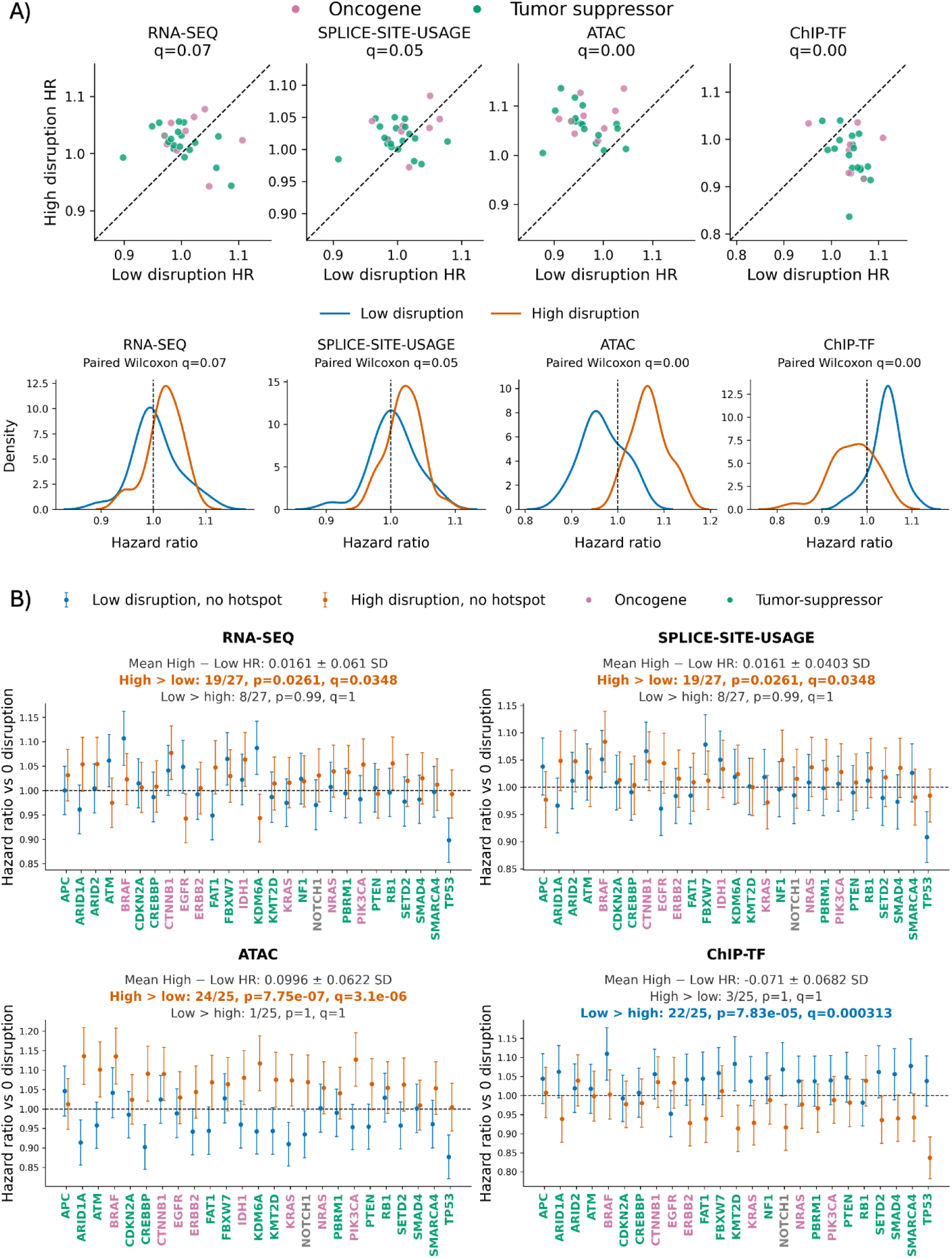
Predicted regulatory disruption from passenger mutations is associated with survival. **(A)** Gene-level comparison of hazard ratios for high-versus low-disruption groups across AlphaGenome modalities. Each point represents a gene, with hazard ratios estimated relative to the zero-disruption/no-hotspot reference group. Dashed lines indicate equal hazard between high- and low-disruption groups. One-sided paired Wilcoxon tests were used to test directional differences across genes, with q-values shown for each modality. **(B)** Density distributions of gene-level hazard ratios for low- and high-disruption groups across modalities. **(C)** Gene-specific hazard ratios for low- and high-disruption groups across RNA-seq, splice-site usage, ATAC, and ChIP-TF modalities. Points show hazard ratios relative to the zero-disruption/no-hotspot reference group, with error bars indicating standard deviation. Text annotations summarize the mean paired difference between high- and low-disruption hazard ratios with a binomial-test p-value and BH-adjusted q-value; significant directional enrichment (q < 0.05) is highlighted. Gene labels are colored by classification (orange, oncogene; blue, tumor suppressor; grey, context-dependent).

Higher predicted disruption was associated with worse survival for several AlphaGenome modalities. In the paired gene-level comparison, ATAC showed the strongest and most consistent pattern: high-disruption hazard ratios exceeded low-disruption hazard ratios in 24 of 25 genes (binomial *q* = 3.1*10^⁻6^), with a mean paired HR difference of 0.0996 ± 0.0622 SD (one-sided paired Wilcoxon *q*< 0.001; **Figure 5A-B**). RNA-seq and splice-site usage showed weaker but concordant effects, with high-disruption hazard ratios exceeding low-disruption hazard ratios in 19 of 27 genes (binomial *q* = 0.0348) for each modality (RNA-seq: mean paired HR difference = 0.0161 ± 0.061 SD, one-sided paired Wilcoxon *q* = 0.07; Splice-site usage: mean paired HR difference = 0.0161 ± 0.0403 SD, one-sided paired Wilcoxon *q* = 0.05; **Figure 5A-B**). These associations held after TMB residualization and cancer-type adjustment, indicating that passenger-mutation-driven regulatory disruption, particularly of chromatin accessibility, carries prognostic signals beyond mutational burden and known driver events. Not all modalities behaved concordantly. For ChIP-TF, the direction was reversed, and the low-disruption group carried higher hazard in 22 of 25 genes (binomial *q* = 0.0003) with a mean paired HR difference of 0.071 ± 0.0682 SD (one-sided paired Wilcoxon *q* < 0.001; **Figure 5A-B**). This suggests that predicted transcription factor binding disruption may not represent generic deleterious gene damage. Instead, ChIP-TF disruption may reflect loss of specific TF-binding programs, including those associated with tumor growth or aggressiveness.

Together, these results indicate that regulatory disruption from passenger mutations, independent of mutational burden, is associated with patient outcome in a modality-specific manner.

### Gene-level disruption is associated with treatment-specific outcomes

Finally, we asked whether AlphaGenome-derived gene disruption scores were associated with survival among patients receiving different classes of therapy in an independent, treatment-annotated cohort. We used the POG570 dataset (Pleasance et al. 2020), which includes whole-genome sequencing, clinical outcomes, and treatment exposure data. Individual drugs were grouped into broad treatment classes based on their primary mechanism of action, including DNA damage inducers, DNA synthesis inhibitors, antimetabolites, hormone therapies, kinase inhibitors, mitotic inhibitors, and growth hormone inhibitors. Because sample size varied across cancer types and treatment groups, we restricted this analysis to breast cancer (BRCA, n=144) and colorectal cancer (COLO, n=87), the two cohorts with the largest numbers of treated patients. Within each cancer type, we selected recurrent driver genes and fit multivariable Cox proportional-hazards models within treatment-class subgroups. Models included AlphaGenome-derived gene disruption scores together with clinical covariates, genome-wide mutational burden, and indicators for co-occurring treatment exposures (**Figure 6**).

**Figure 6:**
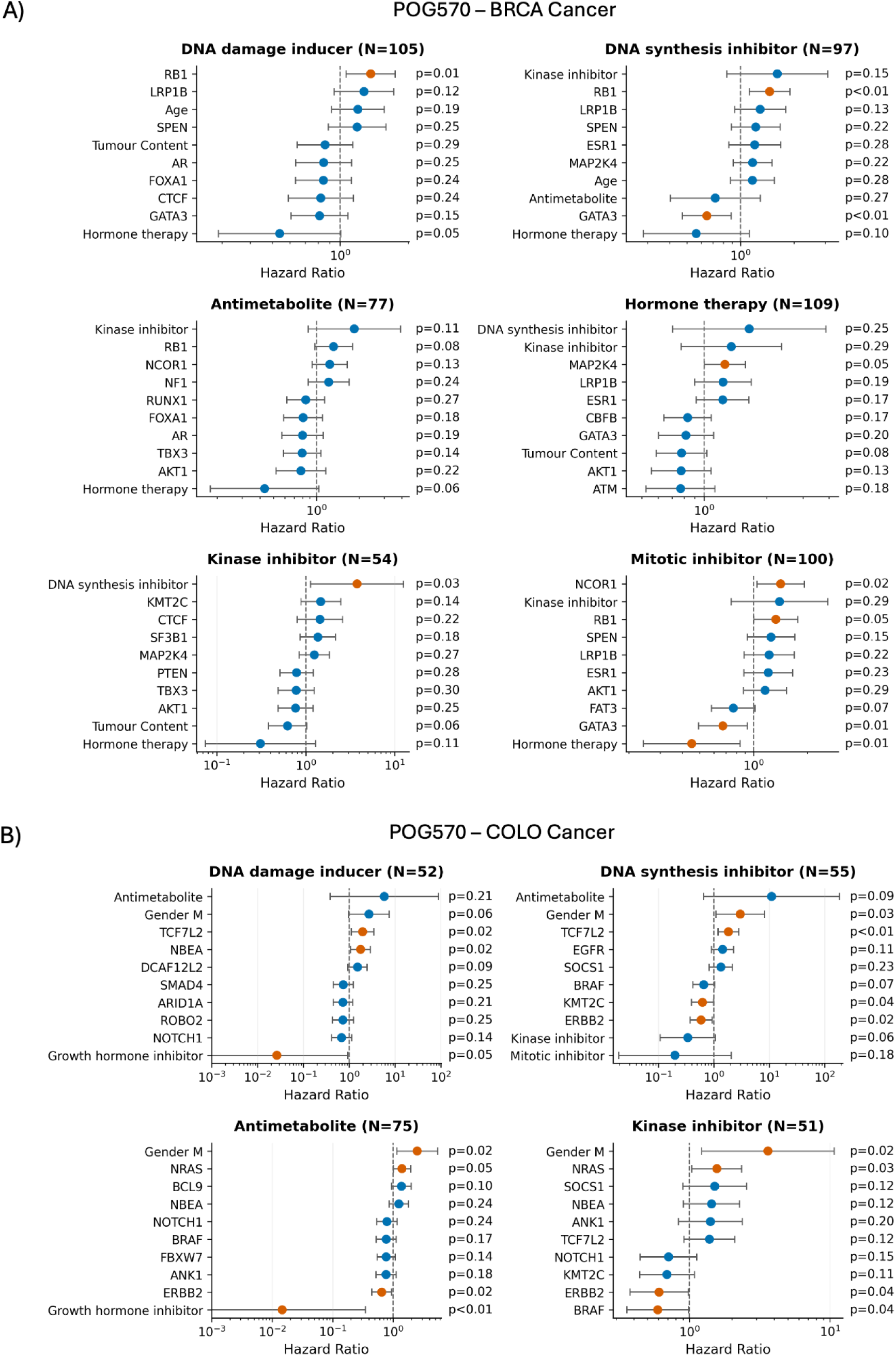
AlphaGenome-derived gene disruption scores are associated with treatment-specific survival outcomes in cancer patients. Multivariate Cox proportional hazards forest plots showing hazard ratios (HR) for the top 10 covariates by p-values, including AlphaGenome-derived gene disruption scores and clinical covariates across treatment class subgroups in two cancer cohorts from the POG570 dataset. (A) BRCA (breast cancer) cohort across six treatment classes. (B) COLO (colorectal cancer) cohort across four treatment classes. Each point represents the estimated HR with 95% confidence intervals on a log scale. Orange points indicate statistically significant associations (p<0.05); blue points indicate nominally non-significant associations. Gene features reflect patient-level AlphaGenome perturbation scores for driver genes, adjusted for co-occurring treatment exposures, age, gender, and genome-wide tumour mutational burden. Dashed vertical line indicates HR=1.

Gene-level disruption scores repeatedly ranked among the top covariates in treatment-specific survival models, alongside established clinical and treatment variables. In BRCA, *RB1* disruption was associated with worse survival among patients treated with DNA damage-inducing agents (HR=1.36, p=0.01) and DNA synthesis inhibitors (HR=1.46, p<0.01). Additional treatment-specific associations included *NCOR1* disruption in the mitotic inhibitor subgroup (HR=1.42, p=0.02) and *GATA3* disruption in both DNA synthesis inhibitor-treated patients (HR=0.65, p<0.01) and mitotic inhibitor-treated patients (HR=0.67, p=0.01). Several clinical or co-treatment variables also showed strong associations, including hormone therapy exposure in BRCA patients treated with DNA damage inducers, antimetabolites, and mitotic inhibitors, highlighting the importance of accounting for concurrent therapies when evaluating gene-disruption effects (**Figure 6A**).

In COLO, AlphaGenome-derived disruption scores also appeared among the strongest treatment-specific covariates. *TCF7L2* disruption was associated with survival among patients treated with DNA damage-inducing agents (HR=1.97, p=0.02) and DNA synthesis inhibitors (HR=1.84, p<0.01), consistent with the relevance of Wnt-pathway disruption in colorectal cancer. *ERBB2* disruption was associated with outcome in patients receiving DNA synthesis inhibitors (HR=0.59, p=0.02), antimetabolites (HR=0.65, p=0.02), and kinase inhibitors (HR=0.61, p=0.04). *BRAF* disruption was also associated with improved survival in the kinase inhibitor subgroup (HR=0.59, p=0.04), while *NRAS* disruption was associated with worse survival in antimetabolite- and kinase inhibitor-treated patients (HR=1.4, p=0.05; HR=1.56, p=0.03; **Figure 6B**).

Together, these results suggest that AlphaGenome-derived disruption scores capture treatment-context-dependent prognostic information in an independent cohort. The recurrence of gene-level disruption features among top covariates across treatment classes supports the idea that regulatory disruption profiles may complement conventional clinical variables and mutation annotations when modeling therapy-associated outcomes. Because subgroup sizes were modest and multiple treatment classes were evaluated, these associations should be interpreted as exploratory and require validation in larger treatment-annotated cohorts.

## Discussion

Here, we applied the genomic foundation model AlphaGenome (Avsec et al. 2026) to quantify the regulatory disruption associated with somatic mutations in cancer genes, showing that these disruption scores capture insights into cancer development and patient outcome. Disruption profiles distinguished cancer genes and recapitulated tissue of origin, cancer type, and microsatellite-instability status; carried MSI-relevant signal that persisted after residualizing on tumour mutational burden; were associated with survival among patients whose disruption arose solely from passenger mutations; and related to treatment-specific outcomes in an independent cohort. Together, these results support a view of cancer-gene perturbation as a graded, multi-modal property rather than a binary driver/non-driver distinction (Nayak et al. 2026; Rheinbay et al. 2020; Martincorena et al. 2018), and suggest that sequence-to-function models can surface functional consequences of somatic variation that coding-centric and burden-based analyses do not capture.

The contrast between AlphaMissense (Cheng et al. 2023) and AlphaGenome is particularly informative because the two models interrogate different axes of variant effect. Recurrent hotspots showed substantially larger predicted protein-level effects, consistent with the strong selective pressure that makes particular coding substitutions recur across tumours (Chang et al. 2016). In contrast, AlphaGenome-predicted regulatory effects were more broadly distributed among variants classified as passengers. This result suggests that recurrence-based and protein-centric prioritization preferentially captures one class of functional consequence, while dispersed somatic variation can perturb transcriptional, chromatin, and RNA-processing programs.

Two features of these findings are worth emphasizing. First, the survival and MSI associations were established on TMB-residualized disruption, so by construction they are not explained by mutational burden, which is the most obvious confounder for any mutation-derived feature. Second, the survival analysis was deliberately restricted to patients without recurrent hotspot mutations, meaning the signal derives from passenger somatic mutations that conventional driver analysis would discard. Such variants need not be functionally neutral, with cohort-level analyses revealing weak positive selection outside canonical driver events (Rheinbay et al. 2020; Kumar et al. 2020). That such variants carry prognostic information is consistent with the hypothesis that somatic mutations can accumulate to produce partial, functionally consequential disruption of cancer genes, extending prior evidence for cumulative gene inactivation (Nayak et al. 2026).

The modality-specificity of these associations is itself informative. Chromatin-accessibility disruption (ATAC) gave the strongest and most consistent survival signal, whereas expression (RNA-seq), transcription factor binding (ChIP-TF), and splicing modalities were moderate. This suggests that the prognostically relevant consequences of passenger mutation may act substantially through regulatory, in addition to protein-coding channels.

Future work will focus on validating predicted disruption against functional and expression data from the same tumours, extending the treatment-response analysis to larger annotated cohorts, and testing whether disruption scores add predictive value in formal, prospectively defined survival and response models beyond the association analyses presented here. More broadly, our results motivate treating cancer-gene function as a continuous, multi-dimensional quantity, and suggest that sequence-model–derived regulatory disruption is a promising, if still preliminary, lens on the functional landscape of somatic variation.

## Methods

### Datasets

Whole-genome somatic variant and clinical data were obtained for 8,800 patients across 33 cancer types from The Cancer Genome Atlas (TCGA) dataset. The dataset is controlled-access and is available upon dbGaP authorization under accession phs000178.v11.p8, and can be downloaded from the ICGC Bionimbus Protected Data Cloud after obtaining access. Survival data were collected from the TCGA Pan-Cancer Clinical Data Resource (TCGA-CDR) (Liu et al. 2018) supplemental data file TCGA-CDR-Supplemental Table S1.xlsx available at https://gdc.cancer.gov/about-data/publications/PanCan-Clinical-2018. The independent POG570 cohort (Pleasance et al. 2020) was used for treatment-associated survival analyses, and the dataset was obtained from https://www.bcgsc.ca/downloads/POG570.

### Cancer Genes and Hotspot annotation

Cancer genes were selected from a predefined cancer-gene set from IntOGen (Gonzalez-Perez et al. 2013). For the pancancer cohort, we use a set of 31 genes that mutated in at least 1% of the patients in the pancancer cohort. For cancer-type-specific gene sets, we use the top 30 genes mutated in most patients in that cohort.

Mutation hotspots were annotated using the Cancer mutation hotspots database (v2) available at https://www.cancerhotspots.org/files/hotspots_v2.xls based on the cancer type of patients. Variants present in the hotspot set were classified as hotspot mutations, while the remaining somatic variants were classified as passengers for the analyses presented here. Files mapping hotspot mutations to the TCGA cohort are available as source data.

### Variant-level scoring for hotspots and passengers

For each single-nucleotide variant, predicted protein-coding effects for missense variants were obtained using AlphaMissense (Cheng et al. 2023). AlphaGenome (Avsec et al. 2026) was used to quantify regulatory effects for all somatic variants assigned to cancer genes. AlphaGenome was run with a fixed 100 kb input sequence length. For each variant, the model input interval was generated by resizing the variant’s reference interval to 100 kb, thereby centering the sequence window on the variant according to AlphaGenome’s interval-resizing implementation. Variants were scored independently using AlphaGenome’s batch variant-scoring API for the human genome.

Regulatory effects were calculated using AlphaGenome’s recommended variant scorers for 7 output types: RNA-seq, CAGE, ATAC-seq, DNase-seq, histone ChIP-seq, transcription-factor ChIP-seq, and splice-site usage. For each variant and output type, AlphaGenome returned effect estimates across the corresponding output type tracks. Non-finite values were removed, and the remaining track-level effects were summarized to obtain the mean signed predicted effects and the mean absolute predicted effect.

### Comparison of hotspot and passenger effects

For the comparison of AlphaMissense and AlphaGenome scores between hotspot and passenger variants, we selected ten pan-cancer driver genes that were mutated in at least 50% of the cancer cohorts. After excluding genes with no mutations in either the hotspot or passenger group, seven genes remained for analysis.

Per-variant AlphaMissense and AlphaGenome effect distributions were compared between hotspot and passenger variants across cancer types and gene classes. For each variant, the AlphaGenome regulatory-effect score was defined as the maximum effect across splice-site usage, RNA-seq, ATAC-seq, and transcription-factor ChIP-seq predictions. These modalities were selected to represent complementary regulatory mechanisms involving RNA processing, transcription, chromatin accessibility, and transcription-factor binding. The maximum was used to capture the strongest predicted regulatory consequence of a variant, because averaging across modalities could attenuate a substantial effect restricted to a single regulatory process. For analyses of effect magnitude, the maximum absolute effect was used, while signed predictions were retained when positive and negative regulatory effects were analyzed separately.

Statistical significance was assessed using two-sided Mann–Whitney U tests, effect sizes were quantified using Cohen’s d, and p-values were adjusted for multiple testing using the Benjamini–Hochberg procedure.

### Gene-level disruption analysis across tissues

To extend the variant-level comparisons to cumulative patient–gene disruption, we analyzed AlphaGenome and AlphaMissense scores across TCGA patient–gene pairs. AlphaGenome analyses used L2_DIFF scores from the four modalities retained in the hotspot–passenger comparison: ATAC-seq, RNA-seq, splice-site usage, and transcription-factor ChIP-seq. Tracks were retained when their strand matched the corresponding gene strand or when they were unstranded, and track-level scores were summarized by the median for each patient, cancer type, gene, and modality.

AlphaGenome modality scores were adjusted for tumor mutational burden, defined as log(1+ total mutations), by dividing each score by TMB. Gene-length effects were then removed by fitting, separately within each patient, tissue, and modality, a linear model of the TMB-adjusted score against log_10_-transformed gene length; model residuals were retained as adjusted scores. These values were z-score normalized across all patient–gene pairs within each modality. For each patient–gene pair, the overall AlphaGenome disruption score was defined as the maximum normalized score across the available modalities, consistent with the variant-level analysis and intended to retain the strongest predicted regulatory consequence. Tissue-gene disruption was summarized as the median patient-level score across samples assigned to each tissue.

For AlphaMissense, the gene-level score was calculated for each patient–gene pair as the sum of AlphaMissense scores across all scored mutations assigned to that gene. Combined scores were restricted to genes represented in the AlphaGenome analysis, z-score normalized across patient–gene pairs, adjusted for TMB and gene length using the same framework, and summarized by the median across patients within each tissue.

For visualization, tissue-gene heatmaps display within-tissue percentile ranks of the summarized disruption scores. In tissues with complete coverage of all four AlphaGenome modalities, the dominant modality for each tissue–gene pair was defined as the modality with the largest summarized normalized disruption score.

### Gene set enrichment analysis

For each cancer type and AlphaGenome modality, we summarized predicted disruption at the gene level by taking the median absolute normalized disruption score across patients. Variant-level AlphaGenome scores were first adjusted for tumor mutation burden and gene length, then z-score normalized within each cancer type and modality. Genes were ranked in descending order of these median absolute disruption scores. We performed preranked gene set enrichment analysis (GSEA) separately for each cancer type, AlphaGenome modality, and Gene Ontology (GO) categories Biological Process and Molecular Function.

Analyses used the Enrichr 2023 GO gene sets (Chen et al. 2013) with a minimum overlap of five genes per term, 1,000 permutations, and normalized enrichment score (NES) as the enrichment statistic. Nominal GSEA p-values were corrected using the Benjamini-Hochberg procedure separately for each combination of cancer type, modality, and GO category. Positive NES values indicate that genes in a GO term tend to have relatively high predicted disruption scores for the given cancer type and modality. Since gene rankings were based on absolute scores, positive enrichment reflects greater predicted disruption magnitude rather than activation, repression, or direction of regulatory change. For cross-cancer heatmaps, we displayed only terms with significant positive enrichment (FDR < 0.05) and significant enrichment in more than one cancer type.

### Tissue- and cancer-type clustering

Using the patient-level AlphaGenome disruption profiles generated above, we evaluated whether predicted regulatory disruption captured tissue- and cancer-specific structure. Disruption profiles included features from all AlphaGenome modalities. Within each modality, tracks were selected according to tissue specificity and, where strand annotations were available, compatibility with the corresponding gene strand.

Continuous features were median-imputed and standardized across all included samples. The resulting patient-level profiles were embedded into two dimensions using t-distributed stochastic neighbor embedding (tSNE) with PCA initialization, an automatically determined learning rate, and a fixed random seed. The five cancer types with the largest numbers of samples were retained for the cancer-type analysis. For tissue-level analysis, cancer types were mapped to their corresponding AlphaGenome tissue categories, and the five tissue categories with the largest numbers of samples were retained. Points were colored by cancer type or tissue type, and group centroids were calculated as the mean t-SNE coordinates and displayed as “X” markers.

### MSI/MSS clustering and classification

To assess whether AlphaGenome disruption profiles captured microsatellite instability (MSI) status, patients from COAD/READ, UCEC, and STAD were assigned high-confidence MSI-H or MSS labels using cohort-specific clinical assays. Discordant or uninformative calls were excluded. For COAD/READ, labels required concordant MANTIS and MSIsensor results, using thresholds of MANTIS > 0.4 and MSIsensor > 3.5 for MSI-H. For UCEC, MSI status required agreement between the five-marker and seven-marker panels. For STAD, MSI status was obtained directly from the available clinical annotation. In both UCEC and STAD, MSI-low tumors were grouped with MSS tumors for consistency. Within each cohort, standardized AlphaGenome profiles were projected using t-SNE as described above, with samples colored by MSI status and group centroids shown as “X” markers.

MSI-H versus MSS classification was evaluated separately within COAD/READ, UCEC, and STAD using stratified cross-validation and three class-balanced logistic-regression models. The first model, M1, used tumor mutational burden, measured as mutations per megabase, as the sole predictor. The second, M2, used AlphaGenome features after removing each feature’s linear association with log-transformed TMB; residualization was learned independently within each training fold and then applied to the corresponding validation fold. The third, M3, used the full AlphaGenome feature set without TMB adjustment.

For all models, missing values were median-imputed, features were max-absolute scaled, and classification was performed using L2-regularized logistic regression with class-balanced weights. Performance was summarized using AUPRC and AUROC. Comparison of the TMB-only and full AlphaGenome models assessed predictive performance relative to the conventional mutational-burden baseline, whereas comparison of the residualized and unadjusted AlphaGenome models quantified the contribution of TMB-related signal to MSI classification.

### Association of gene-disruption with overall survival

To evaluate associations between AlphaGenome-derived disruption and overall survival, we assembled a pan-cancer patient-gene-modality dataset from TCGA patients with available survival and tumor mutational burden data, restricting the analysis to the shared pan-cancer driver-gene set. AlphaGenome disruption scores were measured as described previously, and patient-gene-modality combinations without an available score were excluded. A patient–gene pair was classified as hotspot-mutated when at least one annotated hotspot variant was present in the corresponding gene.

For each gene–modality pair, patients were assigned to four mutually exclusive groups: no hotspot with zero disruption, no hotspot with low non-zero disruption, no hotspot with high non-zero disruption, or hotspot mutation. The zero-disruption group served as the reference. To account for mutation burden when defining disruption strata, non-zero scores were divided by log(1+total mutations) and split at the pan-cancer median for the corresponding gene–modality pair. Patients carrying a hotspot mutation were assigned to the hotspot group irrespective of their disruption score.

A penalized Cox proportional-hazards model was fitted separately for each gene–modality pair, with disruption-group indicators and cancer-type dummy variables included as covariates. Models were fitted only when at least five patients were present in the zero-disruption reference group and at least five overall-survival events were available. Hazard ratios, 95% confidence intervals, and Wald-test p-values were extracted for the low- and high-disruption groups relative to the zero-disruption group. Benjamini–Hochberg q-values were calculated across gene–modality group tests.

For visualization, low- and high-disruption hazard ratios and 95% confidence intervals were plotted for each driver gene and modality. Gene labels were colored according to cancer-gene type (Oncogenes or Tumor suppressor genes), and sample-size labels indicated the combined number of patients in the low- and high-disruption groups. Within each modality, a one-sided binomial test was used to assess whether high-disruption hazard ratios exceeded low-disruption hazard ratios across genes more often than expected by chance. These p-values were adjusted across modalities using the Benjamini–Hochberg procedure.

### Treatment-specific survival analysis in the POG570 cohort

To evaluate whether AlphaGenome-derived gene-disruption scores were associated with treatment-specific clinical outcomes in an independent cohort, we analyzed patients from the treatment-annotated POG570 dataset (Pleasance et al. 2020). Analyses were restricted to the breast cancer (BRCA) and colorectal cancer (COLO) cohorts, which had the largest available sample sizes. Within each cancer type, patients were stratified according to exposure to individual treatment classes. Gene disruption scores were calculated using the same AlphaGenome-based framework described above for the top 30 cancer-specific driver genes by recurrence of mutations.

For each focal treatment class, a single L2-penalized Cox proportional-hazards model for overall survival was fitted using only patients exposed to that treatment. The model jointly included the five retained gene-disruption scores, age, tumour content, log-transformed small-mutation count as TMB, gender, metastatic or recurrent disease status, and binary indicators for exposure to all other treatment classes.

Categorical covariates were one-hot encoded with one reference level omitted. Gene-disruption scores, age, tumour content, and TMB were standardized so that their hazard ratios represented a one-standard-deviation increase. Patients with missing model variables or non-positive survival times were excluded, and models were fitted only for cohorts containing at least 10 patients and three survival events. Hazard ratios, 95% Wald confidence intervals, and nominal Wald p-values were obtained for all model covariates.

## Data availability

TCGA data are available under controlled access and were obtained through dbGaP under accession phs000178.v11.p8 (https://www.ncbi.nlm.nih.gov/projects/gap/cgi-bin/study.cgi?study_id=phs000178.v11.p8). Access requires application and approval through dbGaP. Survival data were obtained from the TCGA Pan-Cancer Clinical Data Resource (TCGA-CDR) (https://gdc.cancer.gov/about-data/publications/PanCan-Clinical-2018). Driver genes were obtained from IntOGen (https://www.intogen.org). The POG570 dataset is publicly available at https://www.bcgsc.ca/downloads/POG570. Hotspot mutations were obtained from the Cancer mutation hotspots database v2 (https://www.cancerhotspots.org/files/hotspots_v2.xls).

## Code availability

Code for data processing, statistical analysis, survival modelling, and figure generation is available at https://github.com/Georgakopoulos-Soares-lab/cancer_non_coding.

## Acknowledgements

The results published here are in whole or part based upon data generated by The Cancer Genome Atlas managed by the NCI and NHGRI. Information about TCGA can be found at http://cancergenome.nih.gov. The TCGA data used for our study can be accessed through dbGaP (phs000178.v11.p8).

This work would not be possible without the participation of our patients and families, the POG team, and the generous support of the BC Cancer Foundation and Genome British Columbia (project B20POG). We also acknowledge contributions towards equipment and infrastructure from Genome Canada and Genome BC (projects 202SEQ, 212SEQ, 12002), Canada Foundation for Innovation (projects 20070, 30981, 30198, 33408), and the BC Knowledge Development Fund. The results published here are in part based upon data generated by the following projects and obtained from dbGaP (http://www.ncbi.nlm.nih.gov/gap): The Cancer Genome Atlas managed by the NCI and NHGRI (http://cancergenome.nih.gov); Genotype-Tissue Expression (GTEx) Project, supported by the Common Fund of the Office of the Director of the National Institutes of Health (https://commonfund.nih.gov/GTEx). All data were generated and are maintained by the BC Cancer Genome Sciences Centre, a part of the Provincial Health Services Authority.

## Author contributions

A.N. and I.G.S. jointly conceived the study. A.N. and T.L. performed data curation, formal analysis, and interpreted the results. A.N. and I.G.S. designed the methodology. A.N. generated the code, statistical analyses, and visualizations. A.N. and I.G.S. wrote the manuscript with input from V.A. I.G.S. acquired funding for the project and provided the supervision.

## Supplementary Figures

**Supplementary Figure 1:**
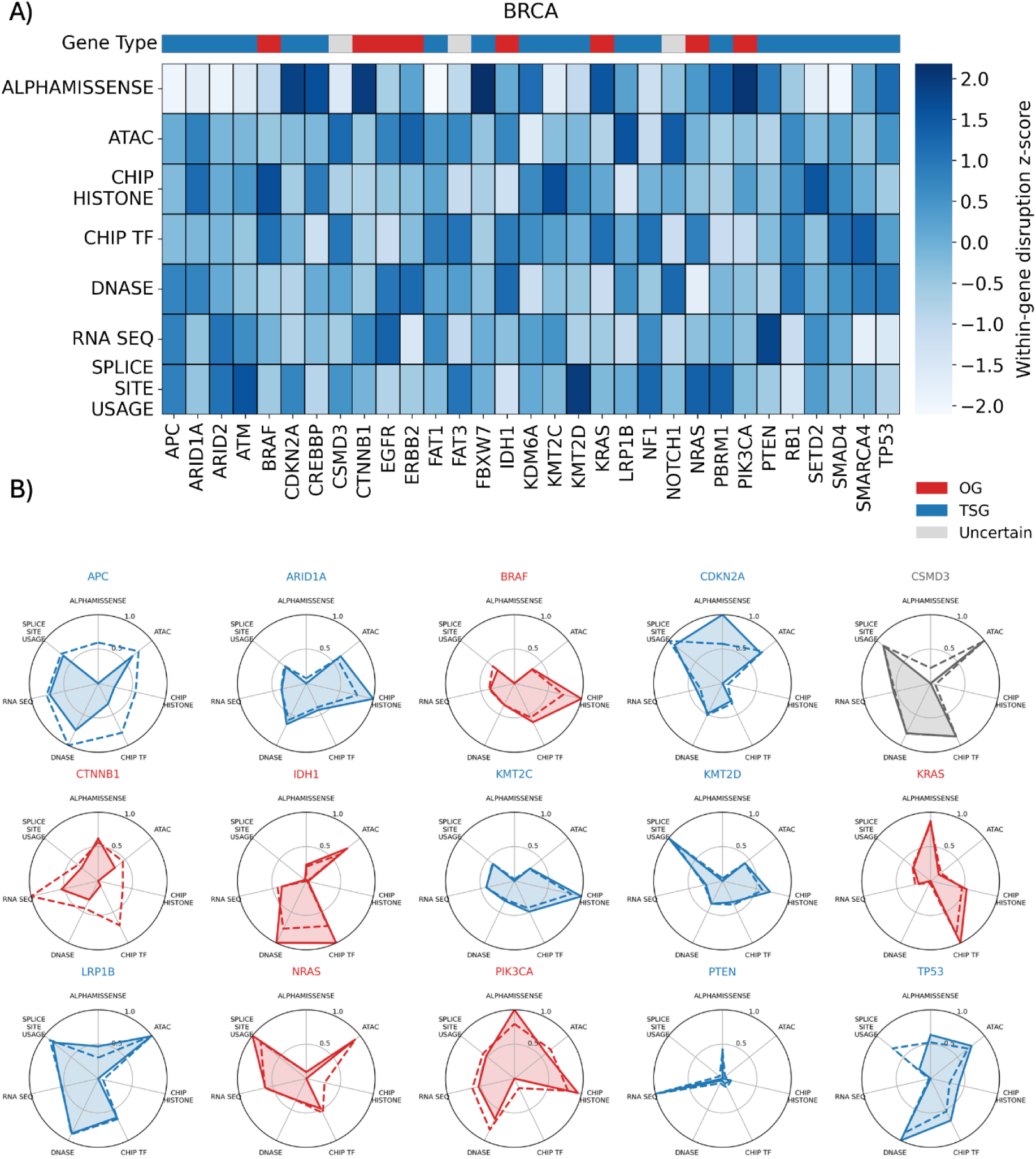
AlphaGenome-inferred regulatory disruption of cancer genes in breast cancer. **(A)** Heatmap of TMB and gene-length-adjusted disruption for cancer genes across AlphaGenome output modalities and AlphaMissense predictions in Breast Cancer (BRCA). Values represent patient-level AlphaGenome and AlphaMissense perturbation scores summarized by gene and modality, adjusted for gene length, and ranked within each modality. Columns are clustered based on gene-level disruption profiles. The gene-type annotation indicates oncogenes and tumor suppressor genes. **(B)** Radial plots for the top 15 driver genes showing mean and median disruption effects across AlphaGenome output modalities and AlphaMissense prediction. Background color indicates gene classification, with red denoting oncogenes, blue denoting tumor suppressor genes.

**Supplementary Figure 2.**
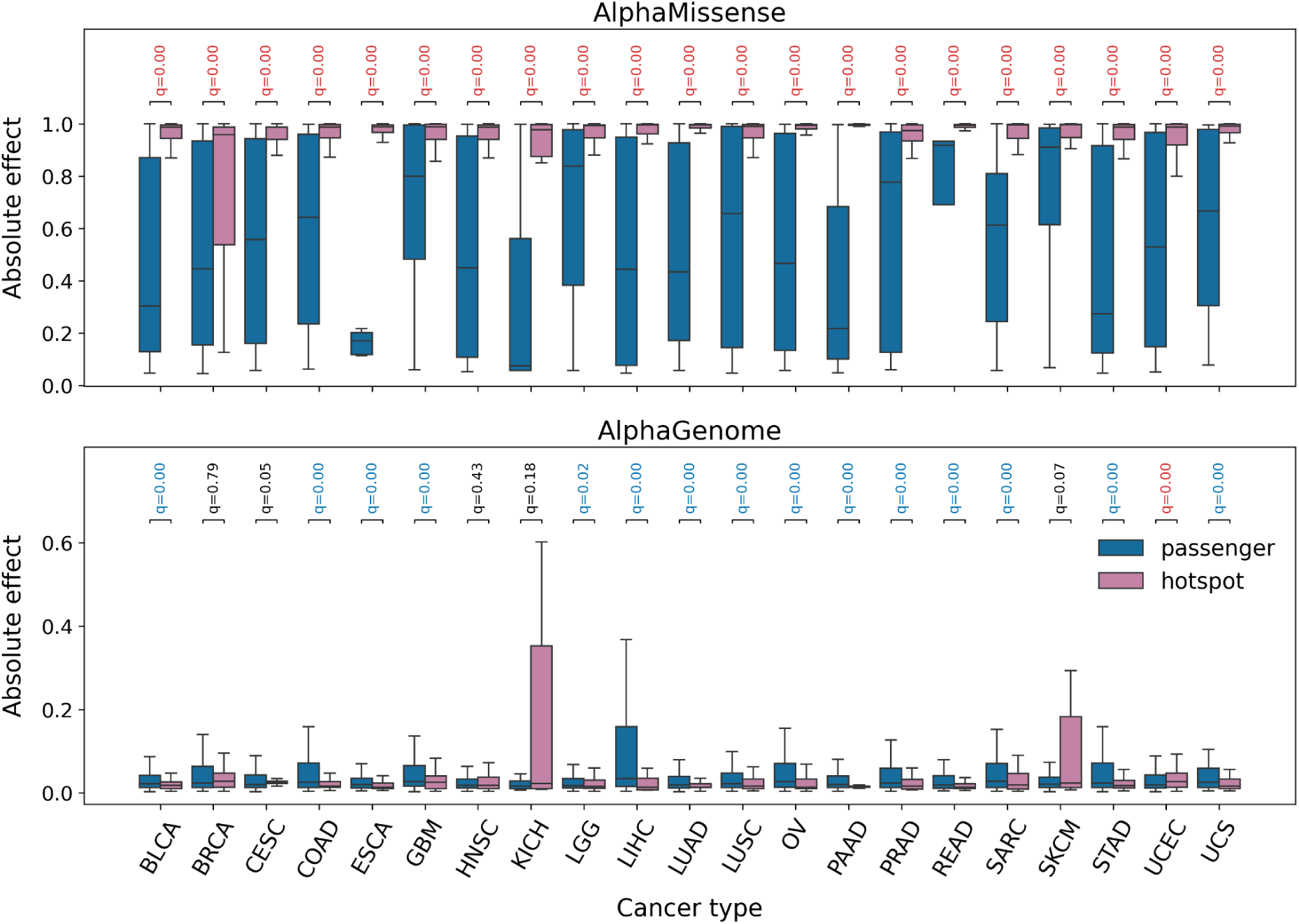
Hotspot and passenger variants show contrasting predicted protein-level and regulatory effects. Distributions of per-variant unadjusted absolute effect scores for passenger and recurrent hotspot variants in top 10 pancancer driver genes, stratified by cancer types. The upper panel shows AlphaMissense scores, and the lower panel shows AlphaGenome-derived regulatory effect scores.

**Supplementary Figure 3:**
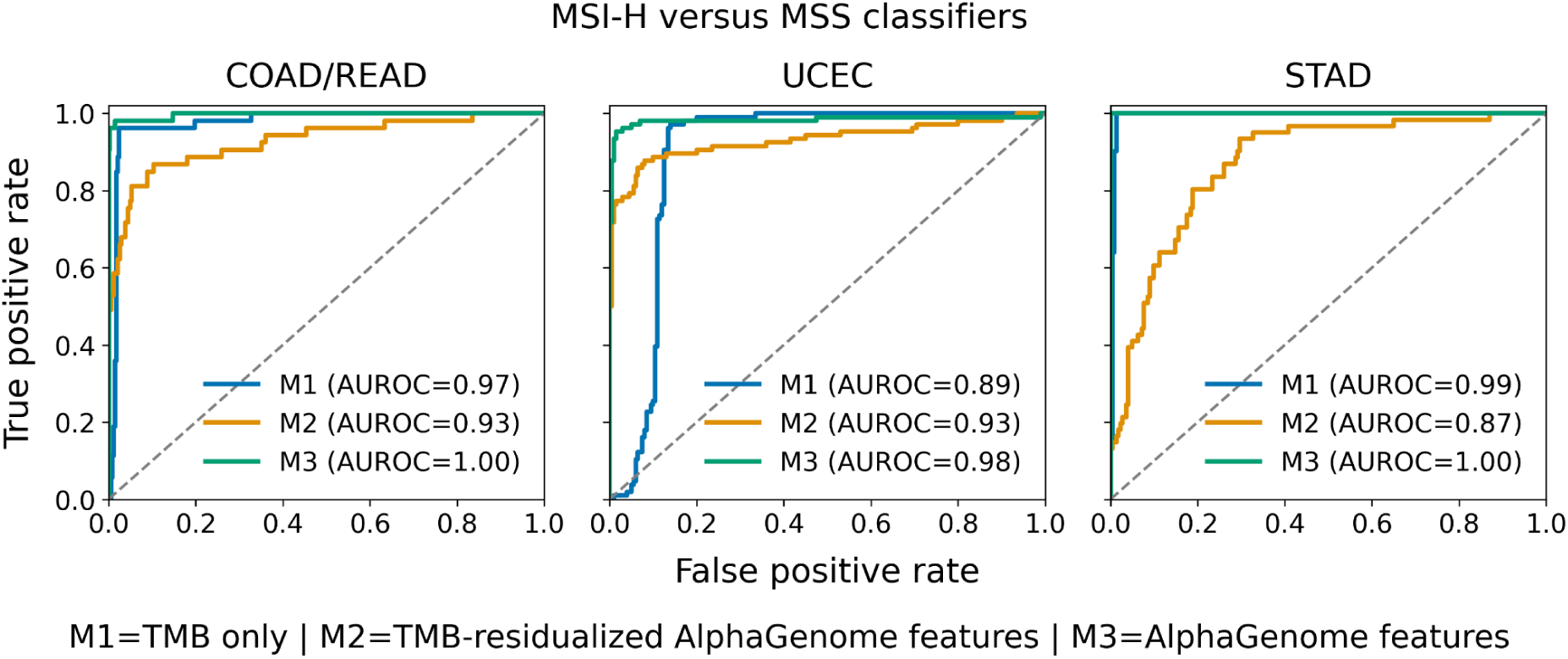
ROC Curves for MSI/MSS classifier models. Models are trained on TMB-only (Model M1), TMB-residualized AlphaGenome features (Model M2), and AlphaGenome features (Model M3).

